# Online information on face masks in Italian and English websites: deficiencies and responsibilities of search engines

**DOI:** 10.1101/2020.10.23.20218271

**Authors:** Shaily Meta, Daria Ghezzi, Alessia Catalani, Tania Vanzolini, Pietro Ghezzi

## Abstract

Countries have major differences in the acceptance of face mask use for the prevention of COVID-19. We analyzed 450 webpages returned by searching the string “are face masks dangerous” in Italy, the UK and the USA using three search engines (Bing, Duckduckgo and Google). The majority (64-79%) were pages from news outlets, with few (2-6%) pages from government and public health agencies. Webpages with a positive stance on masks were more frequent in English (50%) than in Italian (36%), and those with a negative stance were more frequent in Italian (28% vs. 19% in English). Google returned the highest number of mask-positive pages and Duckduckgo the lowest. Google also returned the lowest number of pages mentioning conspiracy theories and Duckduckgo the highest. Webpages in Italian scored lower than those in English in transparency (reporting authors, their credentials and backing the information with references). When issues about the use of face masks were analyzed, mask effectiveness was the most discussed followed by hypercapnia (accumulation of carbon dioxide), contraindication in respiratory disease, and hypoxia, with issues related to their contraindications in mental health conditions and disability mentioned by very few pages. This study suggests that: 1) public health agencies should increase their web presence in providing correct information on face masks; 2) search engines should improve the information quality criteria in their ranking; 3) the public should be more informed on issues related to the use of masks and disabilities, mental health and stigma arising for those people who cannot wear masks.

## Introduction

Face masks are widely recommended, along with hygiene and social distancing, as a measure to prevent the spread of COVID-19 ^1 2^. However, their social acceptance is often problematic with people refusing to use them for several reasons, from health concerns to different social, political and philosophical reasons ^3-5^, and even some leaning towards conspiracy theories. A study on the Twitterverse has also identified a stigma about mask-wearing, which can be seen as a mark for COVID-19 ^6^. On the other hand, there are also contraindications for the use of masks in some health conditions, from respiratory disease to poor mental health or hearing impairment, that are clearly recognized in the WHO guidance ^7^. Issues related to face mask availability, particularly in the initial period of the pandemic, also played a role in the hesitancy about their use ^8^, with health authorities in some countries initially discouraging the use of surgical masks by the general public.

Health authorities have often changed their advice on the use of masks. For instance, the WHO initially did not recommend their use by the general public on 6 April 2020 ^9^, but changed the guidance later during the pandemic, on 5 June 2020 ^7^, with many regional health authorities quickly conforming to the new guidance. There also major differences among different countries, with some governments recommending face masks, some allowing the use of other types of face coverings like scarves (such as in the UK, where reference is often made to “face coverings”) and some regulating their mandatory use differently in closed spaces, outside, on public transports etc.

A lack of a worldwide agreement, and sometimes a lack of alignment to the recommendation by the WHO, along with cultural differences, resulted in marked differences in face mask adoption. Data from a survey on 8-14 June carried out by Imperial College London and YouGov (available at: http://www.coviddatahub.com/) showed that those stating that they “always wore a face mask outside my home” were 85% in Italy, 56% in the USA and 19% in the UK. Conversely, in each country, 1% in Italy, 11% in the USA and 53% in the UK responded that they would not wear a mask.

People often inform themselves about health-related behavior on the Internet, and there is an abundant literature on the importance of online health information in the phenomenon of vaccine hesitancy and anti-vax movements. The COVID-19 pandemic has caused the development of several conspiracy theories about the origin of the virus, the public health measures to deal with it (e.g. face masks, lockdown, isolation, track and tracing apps), as well as the therapeutic strategies against it ^10 11^. These are often heavily politicized and polarized, as shown by studies on what has been called an infodemic ^12^ that includes websites promoting various supplements to “boost immunity”, a topic with an important commercial interest ^13^.

We therefore wondered whether different behaviors in different countries could be related to the type of online information on the potentially negative effects of face masks and their association with conspiracy theories. Using a methodology and a workflow previously used to study anti-vaccine information in different countries ^14 15^, we analyzed the websites returned by searching the string “are face masks dangerous” in Italy’, the UK and the USA. Because different search engines in different countries and languages can differ significantly in the quality of the information they provide on health topics ^14 15^, we used three: Bing, Duckduckgo (a privacy-savvy, no-tracking engine) and Google. We analyzed the first 50 webpages each of them returned in the different localizations in English and Italian. Their content was analyzed in terms of their overall stance towards masks, the presence of conspiracy information and intrinsic transparency features (such as the presence of author, date, references) and the issues discussed in the context of the use of face masks. Finally, we looked at the impact of published guidance on the information online over time.

## Methods

### Search strategy

The search string “are face masks dangerous” (with no question mark) was decided as it was amongst the top suggestions by Google when typing “are face mask”. The corresponding search string in Italian was “le mascherine sono pericolose”. Three search engines were used (Bing, Duckduckgo and Google) using their localized (language and country) version or settings. The search was performed in August 2020. Searches in English were performed in Brighton, UK, setting English language and the USA or the UK as country. Searches in Italian was performed in Urbino, Italy, setting Italy as the country (thus excluding Switzerland). Cookies and previous browsing history were cleared before each search to minimize customization effects.

The first 50 URLs for each search engine result page (SERP) were downloaded to a spreadsheet and subsequently visited for the analysis. All the webpages were archived on the Internet Archive (https://archive.org/web/) on 31 August 2020.

### Analysis

Each webpage was first classified for its typology by two raters (see below for inter-rater agreement). The typologies considered where (based on previous studies on several health topics ^14-17^): blogs, commercial, government, health portals, news, no profit, professional, scientific journals, video and social networks. When an article on a webpage had a date, this was noted as well.

Then the content was analyzed for the mention of conspiracy theories, debunking information and stance about masks. Stance was assigned by two raters trying to answer the question “would a layperson be discouraged from using face masks after visiting this webpage?”. Webpages that did not contain information about face masks were excluded, and the number of excluded webpages for each SERP is reported in the Results section.

Finally, we recorded whether the websites mentioned any negative aspect of face masks and their use (for instance contraindications in children or during exercise, discussing their effectiveness/efficacy or any other issues). When specific words were searched, this was done automatically. Briefly, text corpora were extracted from the webpages to be analysed using WebBootCaT, an online tool for bootstrapping text corpora from the Internet. Then the frequency of specific words was obtained using the corpus analysis software Sketch Engine by Lexical Computing, Brno-Královo Pole, Czechia ^18^. Because some URL does not allow access to robots, not all the webpages were analyzed. Of the 101 unique webpages in Italian, only a sample of 68 (67%) could be analyzed; for the 131 unique URL in English, 91 (69%) could be analyzed.

### Inter-rater reliability

There was a degree of subjectivity in the assignment of website typology and even more in the assessment of the stance towards masks, whether positive, neutral or negative.

The interrater agreement was calculated on a sample of 100 URLs from two raters in Italian (AC and TV) and 89 URLs from two raters in English (SM and DG). Any disagreement was resolved by discussion with a third author (PG). The results for the interrater agreement are reported in the supplementary tables S2-S5.

The interrater agreement for typology in Italian (Table S1) was 90%, with a Kappa of 0.742 (SE 0.073, 95% CI 0.598-0.886); in English (Table S2) it was 92%, with a Kappa of 0.849 (SE 0.054; 95% CI 0.743-0.955). The interrater agreement for the stance on masks in Italian (Table S3) was 79%, with a Kappa of 0.686 (SE 0.061; 95% CI 0.566-0.805); in English (Table S4) it was 66%, with a Kappa of 0.441 (SE 0.081; 95% CI 0.282-0.600). A kappa between 0.41 and 0.60 is considered a “moderate” agreement, between 0.61 and 0.80 “substantial” agreement and between 0.81 and 1.00 an “almost perfect” one ^19^.

The full list of URLs returned in each SERP and their classification, as well as those excluded, are provided as a Supplementary File to allow reanalysis by others.

## Results

### Type of webpages returned

A first analysis looked at the composition of the search results in terms of website typology as described in the Methods section (blogs, commercial, government, health portals, news, no profit, professional, scientific journals, video and social networks). As shown in Table S1, the majority of websites were news outlets (79% on average in Italian, 64% in English-USA and 69% in English-UK), with few others spread across the different typologies. No more than one government website was returned by each search engine (Table S1).

### Content analysis

The content of the webpages was assessed in terms of stance about masks and the presence of conspiracy or debunking information.

Figure 1 shows the percentage of webpages with a positive, neutral or negative stance towards masks for each search (A) and the presence of conspiracy theories or debunking (B).

**Figure 1.**
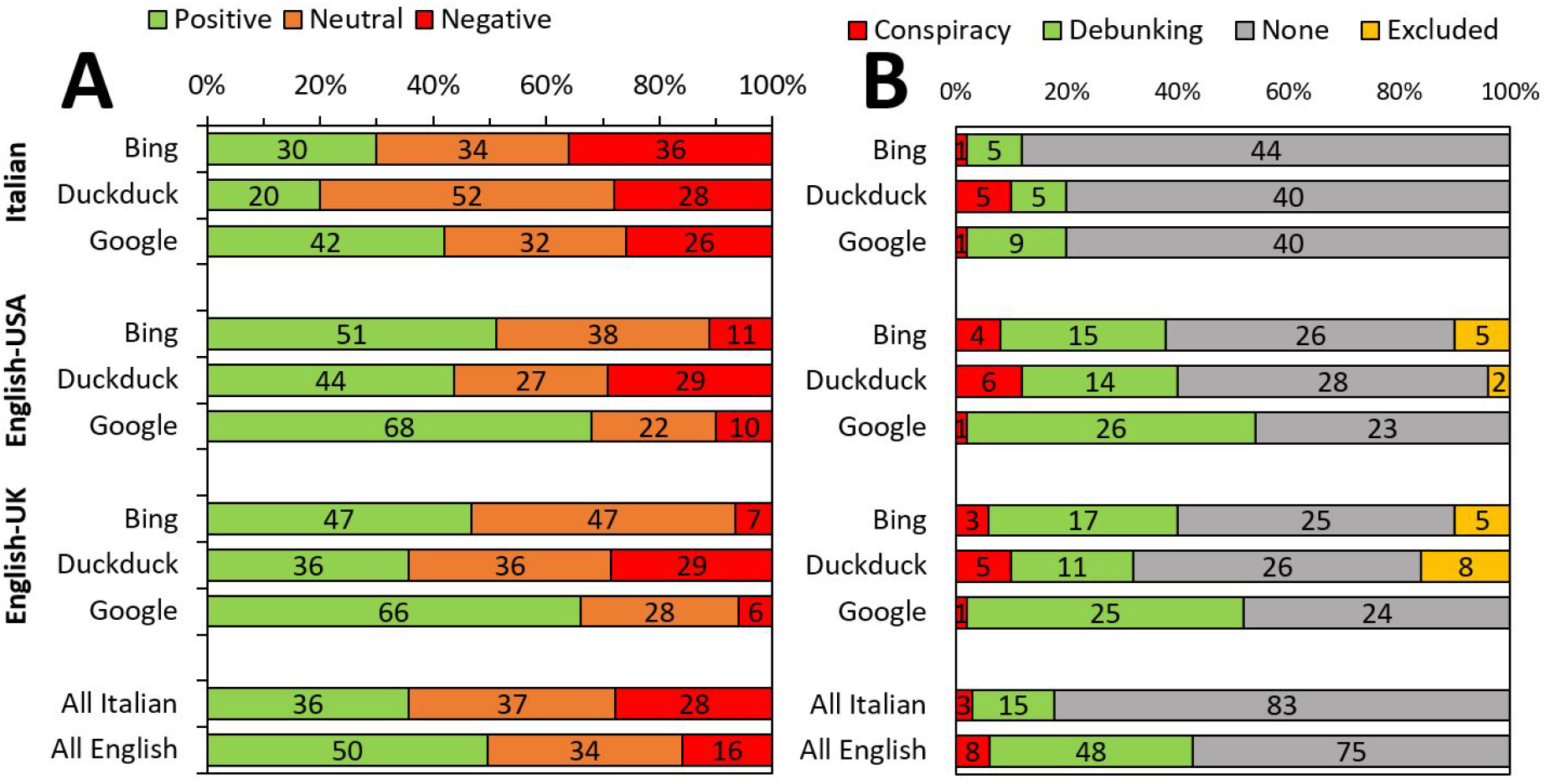
Percentage of webpages in each SERP with different stances on masks (A) and conspiracy or debunking information (B). Panel A. Green, positive; orange, neutral; red, negative. All webpages in each language are all the unique webpages in all the SERPs in that language (Italian, 101; English, 131). Panel B. Conspiracy(red) and debunking (green) information. Number of webpages with conspiracy or debunking information. Total is 50 for each SERP except for Bing USA and Bing UK (n=45), Duckduckgo USA (n=48) and Duckduckgo UK (n=42), as some webpages(in yellow) were excluded for lack of relevance. “All Italian” and “All English” represent all the unique webpages in each language (101 and 131, respectively).

It can be seen that the webpages with a positive stance were more frequent in English (65 of 131, 50%) than in Italian (36 of 101, 36%). There was also a difference in the proportion of negative pages in the three search engines, with Google returning the lowest, followed by Bing and Duckduckgo. We then compared all the unique webpages in Italian with all those in English combining the different SERPs and removing duplicates. For each language, the SERPs (three in Italian and six for English) were combined and duplicates removed, resulting in 101 URLs in Italian and 131 in English. We decided to combine the six SERPs in English because there was a large overlap between the USA and the UK (75 webpages in common, 34 unique for the USA and 22 unique for the UK). As seen from Figure 1A, webpages with a negative stance were more frequent in Italian (28 of 101, 28%) than in English (21 of 131, 16%; P= 0.04 by a Chi-square contingency table). We also analyzed whether the webpages mentioned information related to conspiracy theories or, on the contrary, debunked misinformation. As shown in Figure 1B, the highest number of conspiracy webpages was returned by Duckduckgo and the lowest, on average, by Google, that also had the highest number of debunking webpages.

The ranking of the webpages in the different SERPs is reported in Figure 2, from which it is clear that masks-negative pages are ranked lower by Google in English but not in Italian.

**Figure 2.**
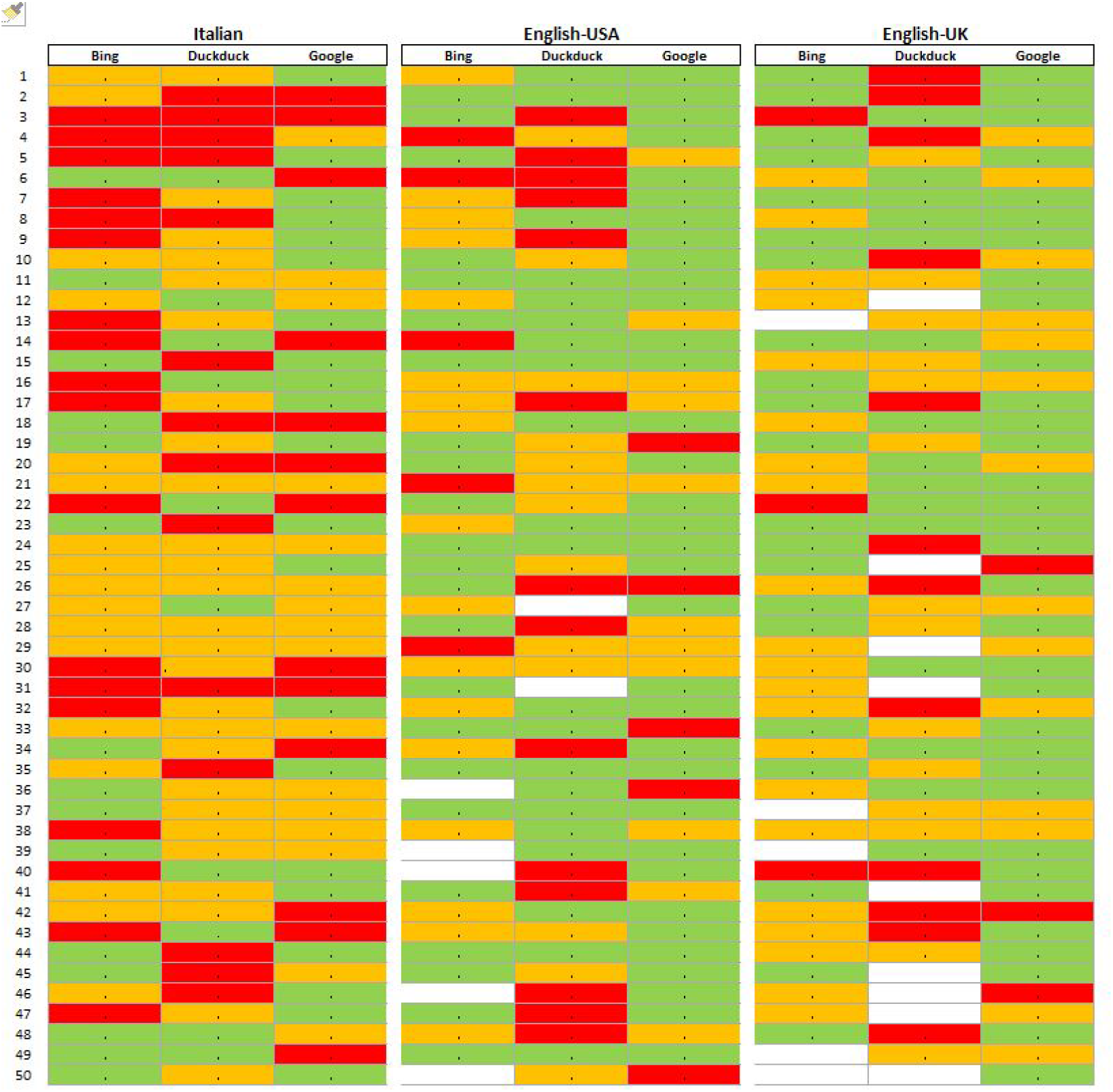
Overall stance on masks in the first 50 webpages in each SERP. Colors indicate the stance: positive (green), neutral (orange) and negative (red). White indicate webpages excluded from the analysis.

### Trustworthiness indicators

We then evaluated the transparency/trustworthiness of webpages by looking at the presence of the following criteria: if the author was given and if so their credentials (e.g. a degree), external reference to back up the information, the date and the ownership of the website. This was done on the 101 webpages in Italian and the 131 in English, as described above.

The results reported in Figure 3 indicate a generally lower presence of trustworthiness indicators in the webpages in Italian. The frequency of each indicator was significantly lower in Italian compared to English (P<0.0001 by Fisher’s test) with the exception of the presence of a date. This deficiency was particularly evident for the presence of the authors’ credentials and of external references. In English, these two indicators were only present in 40-50% of the webpages analyzed.

**Figure 3.**
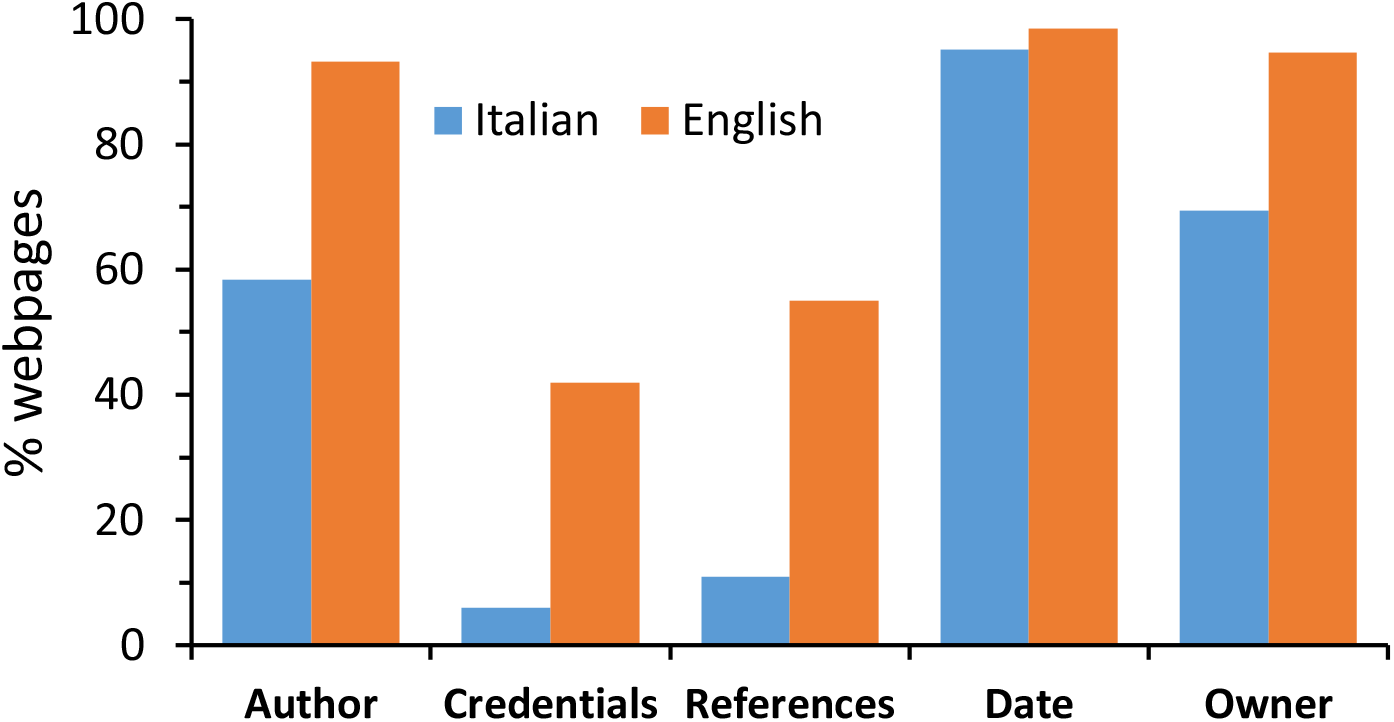
Webpage trustworthiness. Percentage of webpages indicating the presence of transparency indicator (Italian, n=101; English, n=131).

### Variation in the information on masks over time

The guidelines on the use of face masks have changed over time. On 6^th^ April 2020 the WHO recommended their use only by healthcare workers which on the 5^th^ June was extended to the general public following the publication, on 1^st^ June, of a WHO-sponsored study ^1^. In the UK face masks were made compulsory in public transports on 15^th^ May and this was extended to shops on 24^th^ July. In the USA, regulations were very different among states where they became required in shops or public transports between May and July. On 4^th^ April the CDC changed its advice recommending the use of face masks by the general public ^20^. In Italy, following the recommendation of the “Istituto Superiore di Sanità”, they were made compulsory in closed spaces on 26^th^ April, although in Lombardy they were required also outside from 5^th^ April.

Figure 4 shows the frequency of Google searches over the period 1^st^ January – 31^st^ August, using Google trends (https://trends.google.com/) in Italy, the USA and the UK.

**Figure 4.**
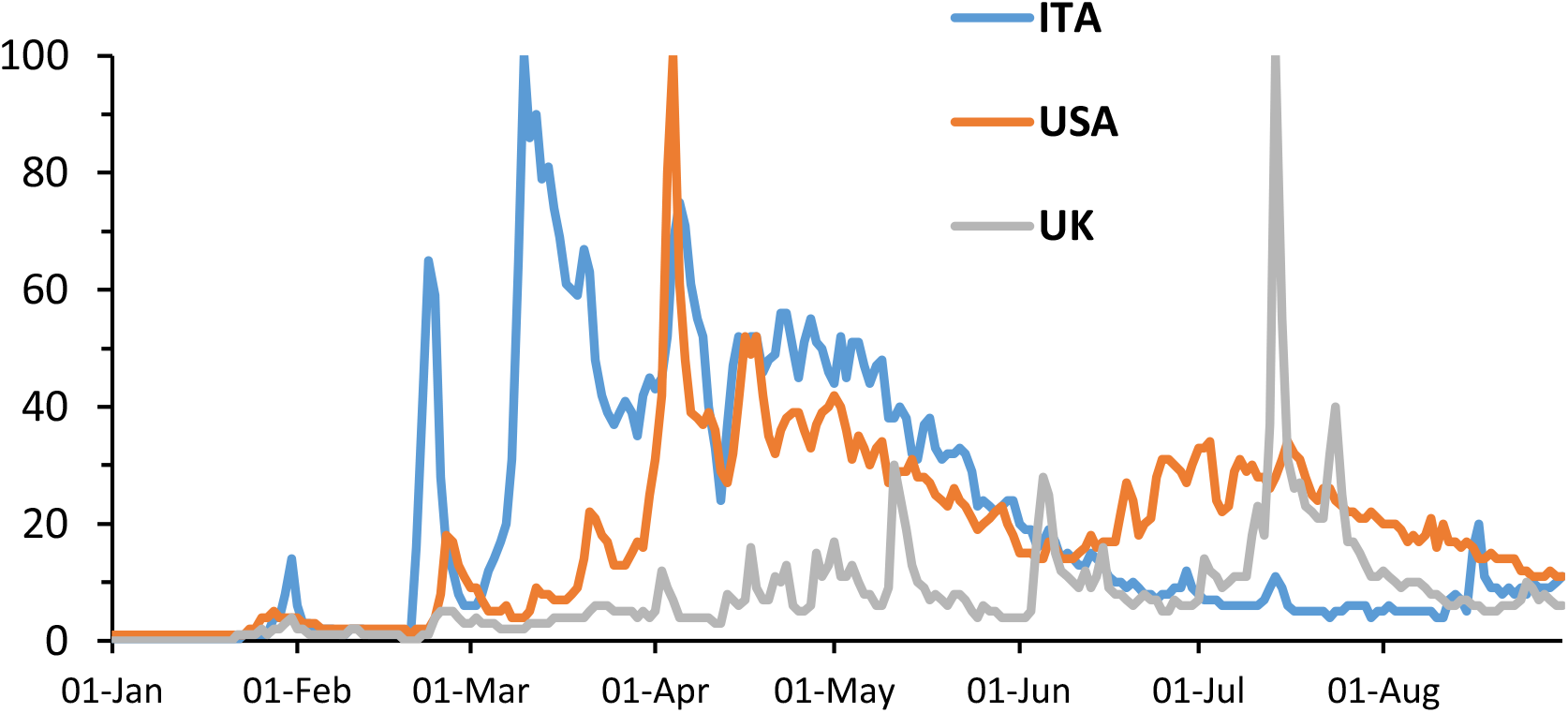
Google searches on face masks. Data represent search interest relative to the highest point on the chart for the given region and time. A value of 100 is the peak popularity for the term in the time range considered. A score of 0 means there were not enough data.

In Italy (searches in Italian in Italy, excluding Switzerland) there were three peaks in the searches on 23^rd^ February, 10^th^ March and 5^th^ April. In the USA the peaks were on 5^th^ April and then in the first half of July. In the UK, the peak was on 15^th^ July. The overall trend followed the introduction of the use of face masks, earlier in Italy, then in the USA and later in the UK.

We also analyzed the stance about masks over time before and after the 1^st^ of June, when the new WHO recommendations were published. This was done on the 101 unique webpages in Italian and 131 in English. The results shown in Figure 5 indicate that there was a trend for a decrease in negative-stance information in Italian and an increase in positive-stance information in English, although these differences were not statistically significant in either language (P=0.11 in Italian, P=0.09 in English; Chi-square test).

**Figure 5.**
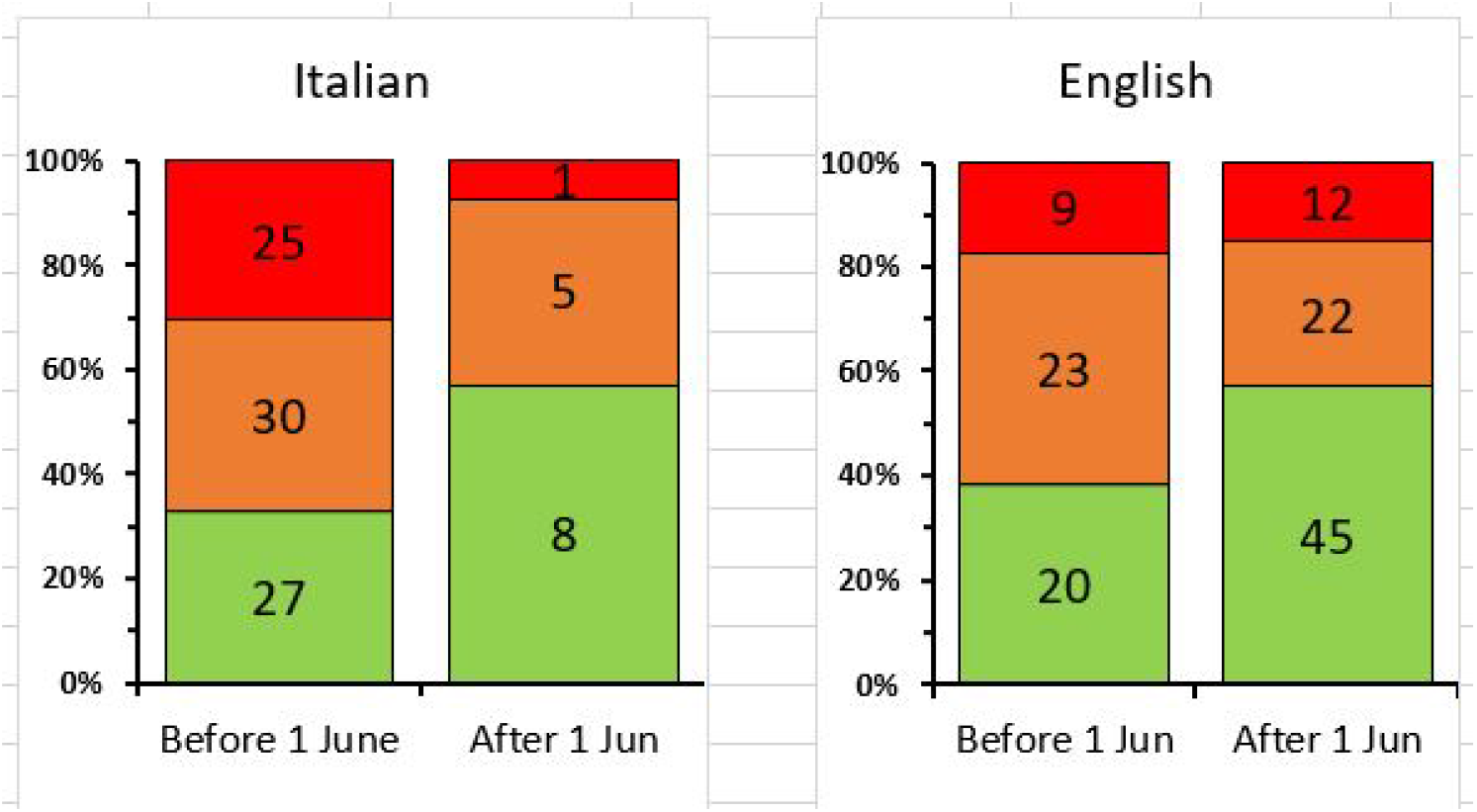
Stance before and after 1^st^ June 2020 in Italian and English.

### Issues associated with the use of masks

We investigated which potential issues associated with the use of masks were mentioned in the 262 unique webpages analyzed (Italian and English combined). As shown in Figure 6, the issue discussed the most often in total was masks’ effectiveness in preventing infection, followed by hypercapnia (due to the accumulation of carbon dioxide), the contraindication in respiratory disease, hypoxia, increased infection (for instance due to re-inhalation), difficulties in using masks properly, their use during exercise, the potential contraindications in children, immunosuppression, the limited availability of masks, and various effects related to mental health or disabilities.

**Figure 6.**
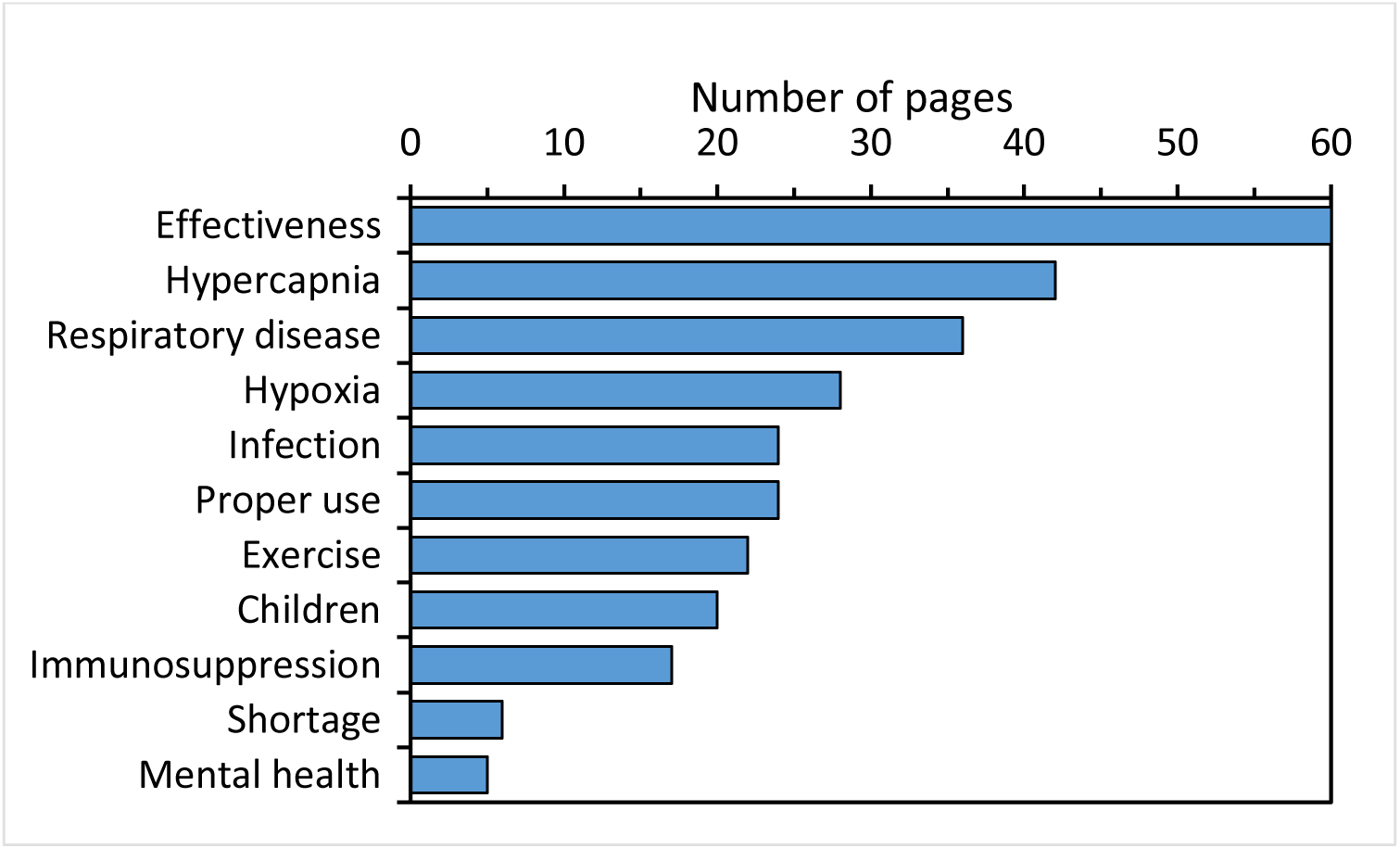
Number of webpages mentioning each type of potential issues in all webpages. Total does not add up to 232 because webpages can mention no issues or multiple issues.

When searching specifically for issues related to hearing impairment (searching the words: hearing, deaf, deafness), we found that this issue was mentioned in 5 of the webpages in English (4%) but only in one of those in Italian (1%). Autism (word) was mentioned only in one webpage in English and in none in Italian. None mentioned stigma that may arise for people for which masks are contraindicated.

## Discussion

A previous study in English and Spanish has analyzed the online information on COVID-19 prevention and found that less than half of the website sample provides recommendation on mask-wearing ^21^. Our study focused on identifying websites that could potentially promote mask hesitancy.

The present study shows that the majority of the websites returned (60-80%) were from news outlets, which was not surprising given the newsworthiness of this topic. However, this first level of analysis on the typology of the websites returned by the search engines highlights a lack of websites from government agencies (between 0% and 2% in the different SERPs), and these websites are supposed to provide high quality information. Professional organizations (universities, hospitals and healthcare organizations), that also usually present high quality information, represented 12%-14% of the pages returned by Google in English, were much less frequent in the SERPs returned by other search engines in English and nearly absent in Italian (including by Google.it with only 2%). This contrasts with the picture we had in previous studies using the same methodology. For example, in a study on vaccine hesitancy in English and Italian, we found between 2% and 6% government websites and 4%-7% professional websites ^14^; in a study on the influenza vaccine in the two languages, government websites returned by Google were between 17% and 42%, and professional ones 4%-19% ^16^. This might be due to the smaller effort made by public health agencies in providing information on face masks compared with vaccination campaigns, but also to the higher ranking given by search engines to news sites.

In general the comparison between the different SERPs confirms the conclusions by our earlier studies ^14 15^ indicating that Google provides information that is more aligned with the scientific consensus, which we interpret as higher information quality, than other search engines, particularly Duckduckgo that also returned the highest number of webpages reporting conspiracy theories and a low number of debunking webpages. However, Google in Italian provides an overall lower quality information than in English. Our findings highlight the ethics responsibilities of search engines that act as gatekeepers of online information.

The other aspect analyzed is the overall stance of the webpages. In total, there was a higher proportion of pages with a negative stance in Italian than in English and more debunking information in English. Italian webpages also fared worse in terms of intrinsic transparency indicators. Of note, four of the trustworthiness criteria analyzed (presence of author, date, references and ownership) are the four components of the most widely used health information quality score, the Journal of the American Medical Association (JAMA) score for websites ^22^. This may seem in contrast with the earlier and higher adoption of face masks in Italy compared to USA or UK. It would be inappropriate to draw a causal link as many other factors, particularly cultural ones, are important in the uptake of face masks. However, this suggests that the presence of information depicting masks in a more negative way (associated with less debunking information) does not have a major impact on behavior of the public.

Because we made some comparison with our previous studies on the online information on vaccines, it is important to note that webpages coded as “negative” on face masks should not be necessarily viewed as “anti-science” in the same way we consider those promoting vaccine hesitancy. In fact, the scientific consensus and guidance on masks changed significantly over time and initially most health authorities, including the WHO, discouraged their use by the general public. In this context, the only webpages that could be compared with anti-vaccine ones are those reporting conspiracy theories but, as shown in Table ***, there were few of them. We are aware of the well reported spread of conspiracy theories on COVID-19 and public health measures in the infosphere ^10 11^ and it is possible that conspiracy theories are more present on social networks.

Another aspect that this research highlighted is the type of issues of face masks that are discussed in websites. Some issues raised frequently are not scientifically based, like the accumulation of CO2 and hypercapnia, hypoxia and the increased risk of infection due, for instance, to re-inhaling the virus or suppressing the immune system. However, other issues are less controversial, like the contraindications in children or people with respiratory complications or psychiatric conditions. Interestingly, few websites mention contraindications in people with some mental health conditions or the consequences of mask use on hearing-impaired persons who lip read ^7^. People that cannot wear masks can be subject to stigma, which could become an increasingly important problem with the increased use of face masks. This problem is particularly evident with issues such as deafness or autism that are mentioned in English webpages but nearly absent in the information in Italian. Of note, a recent study on the Anglophone Twitterverse has not identified any cluster suggesting that discussion around this is probably a small niche compared to politically-motivated anti-masker discourse ^3^ and a recent study highlighted the issue that polarized views can lead to stigma towards both people with and without masks ^23^.

Finally, it is important to note the limitations of the study and its methodology. An obvious limitation is the use of a search string that focused on dangers associated with masks. While this was chosen to address the research question of studying information that could promote mask hesitancy, it clearly resulted in returning more mask-negative information that that present in the whole Web. Also, a limitation is the use of search engines to obtain a sample of the infosphere. Because search engines aim to return high information quality webpages, these could have not returned the lower quality webpages. On the other hand, there are no means of obtaining a random, non-ranked, sample of websites without using search engines, that all have proprietary, non-disclosed, ranking criteria. Also the comparison of two different languages that we attempted might be influenced by cultural differences in the social acceptance of masks and different ranking by the search engines. It should not be forgotten that websites are not the only form of information online. Studies have shown that one quarter of YouTube videos spread misinformation on COVID-19. Because search engines return very few videos, we did not have a sufficient number to try to analyze them separately. Studies searching video sources and social networks for information specifically on face masks should also be done in order to have a more comprehensive picture of the information available online. Social networks, in particular, could play a more important role than websites in promoting conspiracy theories through an “information bubble” effect.

In conclusion, we think this study provides some indication to public health authorities, journalists and healthcare professionals, as well as Internet companies, to correctly inform the public. In particular:

1. The scarce presence of governmental websites suggests that there is room for improving the overall information quality on this topic in the infosphere by a stronger presence of online information by public health agencies but also with news articles citing websites from public health agencies as sources.
2. Search engines such as Duckduckgo, that rightly focus on privacy and no-tracking, should improve the quality of the health information they return and Google could improve their ranking criteria in Italian.
3. The public should be provided with more information on issues related to the use of masks and disabilities, mental health issues and stigma arising for those who cannot wear masks.

## Data Availability

Raw data and URL list available in a Supplementary Online File attached

## SUPPLEMENTARY TABLES

**Table S1.** Typologies of websites returned in the different SERPs.

|  | Italian |  |  | English USA |  |  | English UK |  |  |
| --- | --- | --- | --- | --- | --- | --- | --- | --- | --- |
|  | Bing | Duckduck | Google | Bing | Duckduck | Google | Bing | Duckduck | Google |
| **Blog** | 3 | 3 | 1 | 4 | 6 | 3 | 5 | 5 | 3 |
| **Commercial** | 1 | 1 | 0 | 2 | 1 | 1 | 1 | 1 | 0 |
| **Government** | 1 | 1 | 1 | 1 | 1 | 0 | 0 | 0 | 0 |
| **Health portal** | 1 | 0 | 0 | 1 | 0 | 1 | 1 | 0 | 1 |
| **News** | 38 | 40 | 40 | 33 | 30 | 33 | 36 | 35 | 33 |
| **No profit** | 0 | 0 | 0 | 2 | 1 | 3 | 1 | 0 | 3 |
| **Professional** | 0 | 0 | 1 | 2 | 1 | 6 | 1 | 1 | 7 |
| **Scientific journal** | 0 | 0 | 0 | 0 | 0 | 2 | 0 | 0 | 2 |
| **Video** | 1 | 2 | 1 | 0 | 8 | 1 | 0 | 0 | 1 |
| **Social network** | 1 | 0 | 1 | 0 | 0 | 0 | 0 | 0 | 0 |
| **Excluded** | 0 | 0 | 0 | 5 | 2 | 0 | 5 | 8 | 0 |
| **Other** | 4 | 3 | 5 | 0 | 0 | 0 | 0 | 0 | 0 |

Number of webpages (total, 50).

**Table S2.** Inter-rater agreement for typology (Italian websites)

|  | B | C | G | HP | N | NP | O | P | SJ |
| --- | --- | --- | --- | --- | --- | --- | --- | --- | --- |
| B | 0 | 0 | 0 | 0 | 0 | 0 | 3 | 0 | 0 |
| C | 0 | 1 | 0 | 0 | 0 | 0 | 0 | 0 | 0 |
| G | 0 | 0 | 1 | 0 | 0 | 0 | 0 | 0 | 0 |
| HP | 0 | 0 | 0 | 2 | 0 | 0 | 0 | 0 | 0 |
| N | 0 | 0 | 0 | 0 | 75 | 0 | 1 | 0 | 0 |
| NP | 0 | 0 | 0 | 0 | 0 | 0 | 0 | 0 | 0 |
| O | 0 | 0 | 0 | 0 | 6 | 0 | 12 | 0 | 0 |
| P | 0 | 0 | 0 | 0 | 0 | 0 | 0 | 1 | 0 |
| SJ | 0 | 0 | 0 | 0 | 0 | 0 | 0 | 0 | 0 |

Number of websites assigned to each typology by two raters. Legend: B, blog; C, commercial; G, government; HP, health portal; N, news; NP, no profit; O, other; P, professional; SJ, scientific journal.
Number of observed agreements: 92 ( 90.20% of the observations); Number of agreements expected by chance: 63.2 ( 62.00% of the observations); Kappa= 0.742; SE of kappa = 0.073; 95% confidence interval: From 0.598 to 0.886; Weighted Kappa= 0.630.

**Table S3.** Inter-rater agreement for typology (English websites)

|  | B | C | G | HP | N | NP | O | P | SJ |
| --- | --- | --- | --- | --- | --- | --- | --- | --- | --- |
| B | 6 | 0 | 0 | 0 | 2 | 0 | 0 | 0 | 0 |
| C | 0 | 1 | 0 | 0 | 0 | 0 | 0 | 0 | 0 |
| G | 0 | 0 | 1 | 0 | 0 | 0 | 0 | 0 | 0 |
| HP | 0 | 0 | 0 | 3 | 1 | 0 | 0 | 0 | 0 |
| N | 1 | 0 | 0 | 0 | 58 | 0 | 0 | 0 | 0 |
| NP | 1 | 0 | 0 | 0 | 1 | 5 | 0 | 0 | 0 |
| O | 0 | 0 | 0 | 0 | 0 | 0 | 1 | 0 | 0 |
| P | 0 | 0 | 0 | 0 | 0 | 0 | 1 | 5 | 0 |
| SJ | 0 | 0 | 0 | 0 | 0 | 0 | 0 | 0 | 2 |

Number of websites assigned to each typology by two raters. Legend: B, blog; C, commercial; G, government; HP, health portal; N, news; NP, no profit; O, other; P, professional; SJ, scientific journal.
Number of observed agreements: 82 ( 92.13% of the observations); Number of agreements expected by chance: 42.8 ( 48.06% of the observations); Kappa= 0.849; SE of kappa = 0.054; 95% confidence interval: From 0.743 to 0.955; Weighted Kappa= 0.846.

**Table S4.**
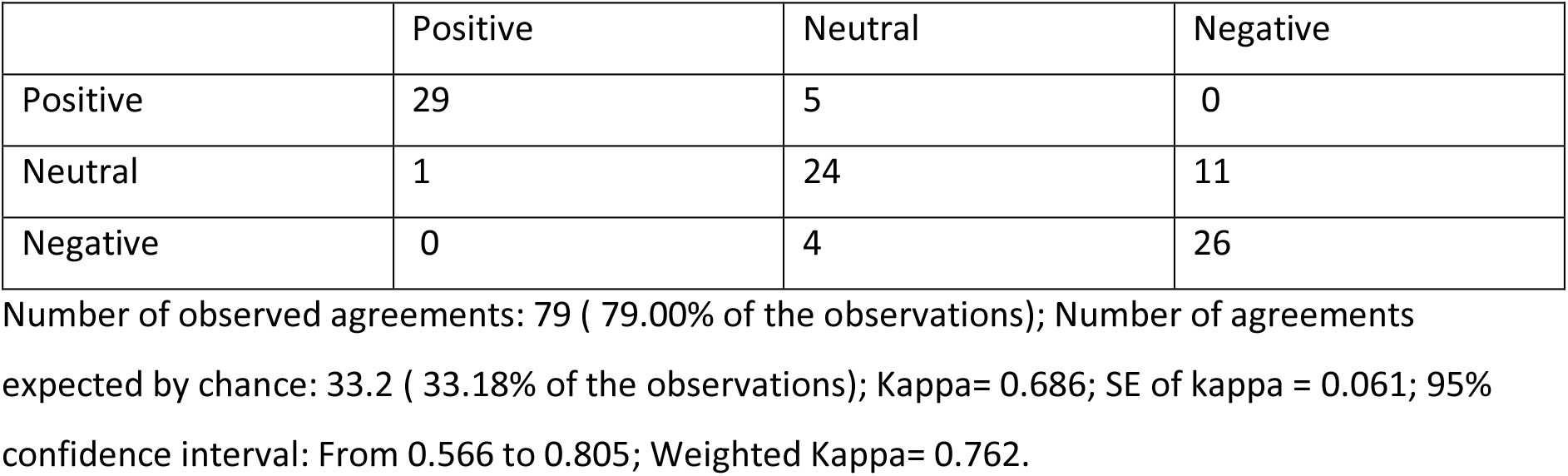
Inter-rater agreement for stance on masks (Italian webpages)

|  | Positive | Neutral | Negative |
| --- | --- | --- | --- |
| Positive | 29 | 5 | 0 |
| Neutral | 1 | 24 | 11 |
| Negative | 0 | 4 | 26 |

Number of observed agreements: 79 ( 79.00% of the observations); Number of agreements expected by chance: 33.2 ( 33.18% of the observations); Kappa= 0.686; SE of kappa = 0.061; 95% confidence interval: From 0.566 to 0.805; Weighted Kappa= 0.762.

**Table S5.** Inter-rater agreement for stance on masks (English webpages)

|  | Positive | Neutral | Negative |
| --- | --- | --- | --- |
| Positive | 34 | 12 | 1 |
| Neutral | 6 | 18 | 5 |
| Negative | 3 | 3 | 7 |

Number of observed agreements: 59 ( 66.29% of the observations); Number of agreements expected by chance: 35.4 ( 39.73% of the observations); Kappa= 0.441; SE of kappa = 0.081; 95% confidence interval: From 0.282 to 0.600; Weighted Kappa= 0.491.

